# Refractive error among school-aged children in Bhutan: A secondary analysis of data from nationwide Bhutan School Sight Survey

**DOI:** 10.1101/2025.08.05.25333029

**Authors:** Indra Prasad Sharma, Kovin S Naidoo, Khathutshelo Percy Mashige, Nor Tshering Lepcha

**Author notes:** **Corresponding author**, [IPS].

## Abstract

To determine the prevalence, demographic distribution, and predictors of refractive error among school-aged children in Bhutan using secondary data from the Bhutan School Sight Survey (BSSS), and to compare these population-level findings with standardised estimates from the Refractive Error Study in Children (RESC). Secondary data from 586 schools, comprising 166,357 school-aged children (5-18 years), were analyzed. As part of the BSSS, vision screening and cycloplegic refraction were conducted following the RESC protocol. Spectacles were provided at no cost to children requiring correction. Data on demographic characteristics (age, gender), school type, school level, and geographic location were analysed using descriptive statistics. Logistic regression was used to identify associated predictors. Findings were compared with previously published RESC data to contextualise observed trends. Visual impairment (VI) affected 12.14% (95% CI: 11.98-12.30) of school-aged children in Bhutan, with higher rates in females (13.16%) than males (11.07%). Refractive error (RE) was the leading cause, accounting for 11.71% of all cases and 96.1% of VI. Myopia was most common (5.78%), followed by astigmatism (5.26%) and hypermetropia (0.67%). Multivariate analysis showed female sex, private school attendance, and urban schooling (Thromdes) as significant RE predictors (p < 0.01). Of 19,234 children diagnosed with RE, 96.2% received spectacles within four months; 3.8% were lost to follow-up. Effective spectacle coverage was 11.5%.In Bhutan, refractive error affects over 10% of school-aged children, with higher prevalence among females, urban students, and those in private schools highlighting equity gaps in eye health. These findings are consistent with prior estimates derived from the Refractive Error Study in Children (RESC) protocol and underscores the need for sustained, targeted school-based eye care. The BSSS also shows that comprehensive screening is feasible and scalable in resource-limited settings.

## Introduction

Uncorrected refractive error (URE) is the most readily treatable form of visual impairment (VI), yet it persists as the leading cause of VI worldwide and a major contributor to preventable vision loss among school-aged children. The prevalence of URE, particularly myopia, has reached epidemic proportions, with the highest burden observed in East and Southeast Asia [1]. Despite the availability of effective corrective interventions, only 36% of individuals worldwide with distance VI attributable to refractive error (RE) have access to appropriate refractive correction [2].

URE has been shown to negatively affect children’s academic achievement, social functioning, and overall developmental outcomes [3]. A pooled analysis of global data estimated the prevalence of URE among individuals under 20 years of age in Southeast Asia to be approximately 2.26 per 1000, highlighting the significant regional burden [4]. Notably, the prevalence of myopia has demonstrated a marked upward trend, with projections indicating that over 740 million children and adolescents worldwide will be affected by 2050 [5]. In response to the growing of URE, the World Health Organization (WHO) launched the SPECS 2030 initiative, which targets a 40% increase in effective refractive error coverage (eREC) by 2030 [6]. However, the realisation of this target necessitates robust, evidence-informed strategies, yet current national-level data remain limited, highlighting the urgent need for comprehensive, population-based survey data [7].

Bhutan has demonstrated a strong commitment to universal health coverage, with eye health integrated into its national public health priorities [8]. Bhutan is also one of the few countries to hold a workshop for WHO SPECS 2030 initiative. Despite this, the country’s mountainous geography and widely dispersed rural population have posed significant challenges to the delivery of pediatric eye care. Prior to 2019, there was a notable absence of systematic data on RE among children. In response, the Ministry of Health, in partnership with the Ministry of Education and international stakeholders, implemented the Bhutan School Sight Survey (BSSS) in 2019; a comprehensive, nationwide, school-based RE screening initiative. . The BSSS attained a high school coverage rate of 99.5%, screening a total of 164,365 children, which represented 98.8% of the national school-age population. A total of 16,376 spectacles were dispensed free of charge as part of this initiative [9].

Despite the successful implementation of the BSSS, comprehensive analysis of the dataset was delayed due to the reallocation of national priorities following the onset of the COVID-19 pandemic, leaving the dataset largely unexamined. A component of the BSSS, using the Refractive Error in School Children (RESC) protocol, was published [10]; however, further integrative analysis combining the RESC-based findings with the broader BSSS data was not undertaken. This secondary analysis of the BSSS data seeks to estimate the national prevalence of RE among children, examine its distribution across demographic and geographic subgroups, and identify key predictors. Additionally, the analysis aims to compare RESC-derived estimates and census-level findings. The resulting findings are expected to enhance the validation and interpretation of population-based screening methodologies.

Supported by robust, population-level data, the findings of this study are anticipated to inform evidence-based policy formulation for school-based eye health interventions and contribute meaningfully to Bhutan’s advancement towards meeting the WHO eREC target for 2030.

## Methodology

### Study design and setting

This study presents a secondary analysis of the BSSS dataset. The BSSS represents the first nationwide, school-based eye screening program conducted in Bhutan, carried out from March to December 2019. Initiated by the Ministry of Health in collaboration with the Ministry of Education and international partners, the BSSS aimed to screen all school-aged children for RE and provide corrective spectacles as a public health intervention. This secondary analysis seeks to investigate previously unreported associations and generate stratified prevalence estimates not addressed in the initial RESC-based publication [10]. The BSSS adhered to the standardized RESC Protocol, facilitating comparability with international studies. The principal investigator of the current analysis also served as the lead investigator for the BSSS program.

### Study Population and Sampling

The BSSS encompassed 586 of 589 schools across all 20 Dzongkhags (districts) and four Thromdes (municipalities) in Bhutan, achieving a coverage of 99.5%. A total of 164,365 children aged 5 to 18 years, representing 94.3% of the school-enrolled population in Bhutan, were examined. The analysis included all children enrolled in schools during the study period who underwent visual acuity screening and refraction as part of the survey. Sampling procedures were not applied, as the BSSS aimed to achieve universal school coverage.

### Protocol for BSSS

#### Survey team

Six teams were deployed to conduct vision screening, refraction, spectacle dispensing, and data collection. Each team comprised six optometrists and eight ophthalmic technicians. The ophthalmic technicians were responsible for performing vision screenings and recording data, whereas the optometrists conducted clinical assessments and diagnoses.

#### Training and quality assurance

The survey team completed a two-day hands-on training focused on the examination procedures, data collection, and quality assurance protocols. A pilot study was subsequently conducted to familiarize team members with all aspects of the study protocol. Additionally, the PI and study investigators provided continuous supervision and monitoring fieldwork throughout the survey to ensure strict adherence to the protocol.

#### Vision screening and refraction procedure

The examination protocol followed that described in the previous study [10]. In brief, all participants underwent an initial assessment of uncorrected visual acuity (UCVA) at 6 meters using a Tumbling E chart, administered at temporary stations established within each school. Students with a UCVA of ≤ 6/12 in either eye were subsequently evaluated by an optometrist for both objective and subjective refraction. Cycloplegic refraction was performed using 1% cyclopentolate and measured with a Plusoptix A12R autorefractometer (Plusoptix GmbH, Nuernberg, Germany), followed by subjective refraction to determine the best-corrected visual acuity (BCVA) [10]. Fundoscopic examination and binocular motor function tests were not conducted. Visual impairment (VI) was attributed to refractive error (RE) if VA ≤ 6/12 in either eye improved to ≥6/9 following subjective refractive correction.

Children diagnosed with RE received prescription spectacles, while those whose VA did not improve to ≥6/9 after correction were referred to the nearest ophthalmology center for further assessment and management.

### Definitions

The BSSS utilized the definitions specified in the RESC protocol. UCVA of 6/9 or better in both eyes was classified as normal vision and, while excluded from refraction consideration, was included in the calculation of visual impairment (VI) prevalence rates. VI was defined as UCVA of ≤ 6/12 in the better eye. URE was defined as a presenting VA of ≤ 6/12 in either eye that improved to ≥ 6/9 following refraction. Myopia was defined as a spherical equivalent (SE) of ≤-0.50 diopters (D), hypermetropia as ≥ +2.00 D, and astigmatism as a cylindrical power of≥ ±0.75 D in either eye.

### Data Sources and Variables

For this secondary analysis, cleaned datasets were obtained from the Primary Eye Care Program, Jigme Dorji Wangchuck National Referral Hospital, Ministry of Health. All data were anonymized and authors could not identify individual participants.The data was obtained on 8th November 2023 after completing all official procedures. Variables included demographic characteristics stratified by gender, school type, school level and school location. Data captured comprised the total number of students with VI and RE, with RE further classified by gender. Spectacle distribution data were sourced from the Primary Eye Care Program. Enrollment records available from the website of the Ministry of Education were used to validate coverage and ensure representativeness of the sample.

### Statistical Analysis

Descriptive statistics were employed to summarize demographic variables and estimate the prevalence of VI and RE, stratified by gender, school type, school location, and geographic region. Bivariate analyses were conducted to examine differences in prevalence across these subgroups. Multivariate logistic regression models were used to identify independent predictors of overall RE and its specific subtypes (myopia, hypermetropia, and astigmatism). Odds ratios (ORs) with corresponding 95% confidence intervals (CIs) were reported, and statistical significance was set at p < 0.05. All statistical analyses were performed using IBM SPSS Statistics (IBM Corp., Armonk, NY, USA), Version 26.0.

### Ethical Considerations

The original BSSS survey received ethical approval from the Research Ethics Board of Health (REBH), Ministry of Health, Bhutan, with additional authorization granted by the Ministry of Education. The survey was conducted as a public health intervention using standardized eye examination protocols. Written informed consent was obtained from school authorities, including principals and teachers, while verbal assent was secured from participating students. The REBH also approved this secondary analysis of anonymized data without requiring re-consent.

## Results

### Survey Coverage

The BSSS encompassed 99.5% of the schools in Bhutan, examining 586 out of 589 schools. A total of 164,365 (98.7%) schoolchildren underwent eye examinations, with a gender distribution of 49% male (n=80,469) and 51% female (n=83,896). Three schools, representing 518 students, were inaccessible due to adverse climatic conditions and logistical challenges. Additionally, 1,427 students were not examined due to absence on the examination day, school withdrawal, or refusal to participate. According to the Population and Housing Census of Bhutan (PHCB 2017), the estimated population of children aged 5 to 18 years was 174,377. Consequently, the BSSS achieved coverage of 94.3% of the target population in this age group, as shown in **Fig 1**.

**Fig 1.**
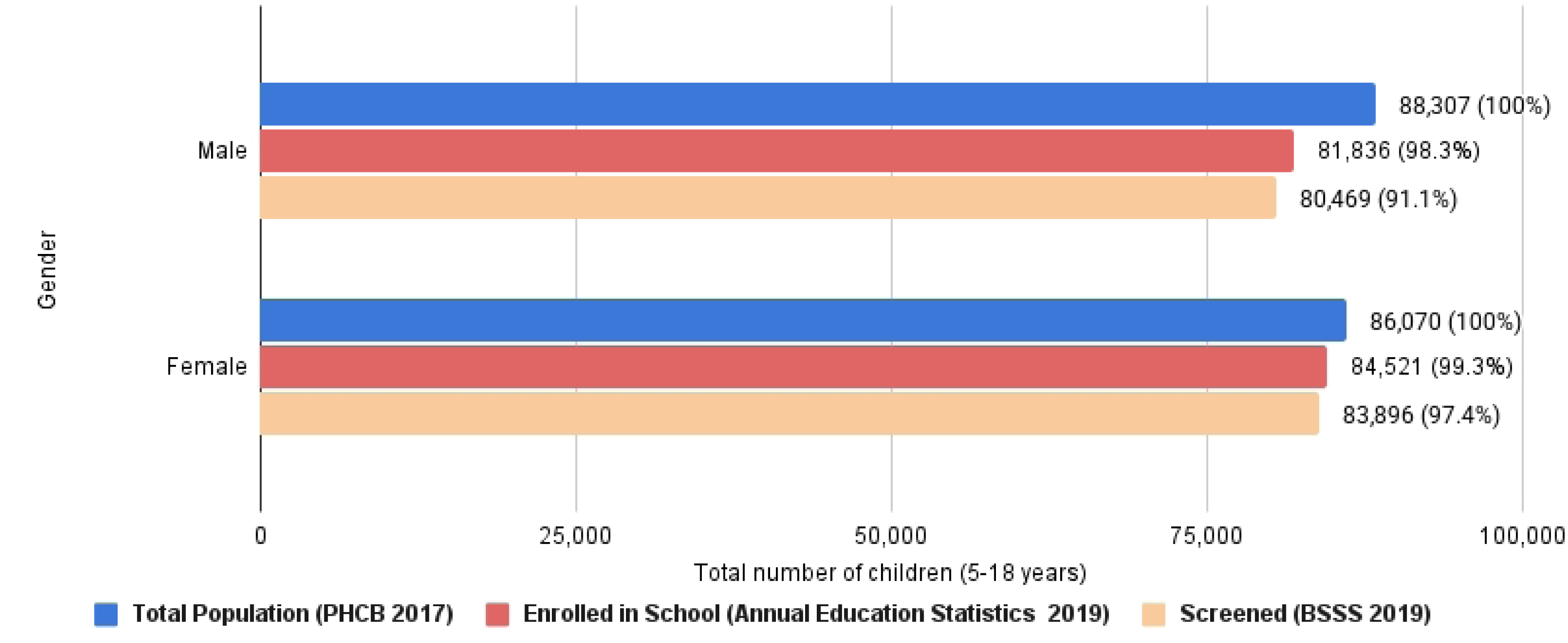
Coverage of the children during the Bhutan School Sight Survey.

### Prevalence of VI and URE

Among the 164,365 school-aged children screened, 144,414 (87.87% 95% CI:87.71% - 88.03%) had normal uncorrected vision (UCVA ≥ 6/9 in both eyes). The overall prevalence of VI was 12.14% (95% CI: 11.98 - 12.30), with a higher prevalence observed in females (13.16%) compared to males (11.07%). URE accounted for 11.71% (95% CI: 11.55 -11.86) of cases and represented 96.1% of all identified VI. A summary of the visual acuity assessment findings is presented in **Table 1**.

**Table 1.**
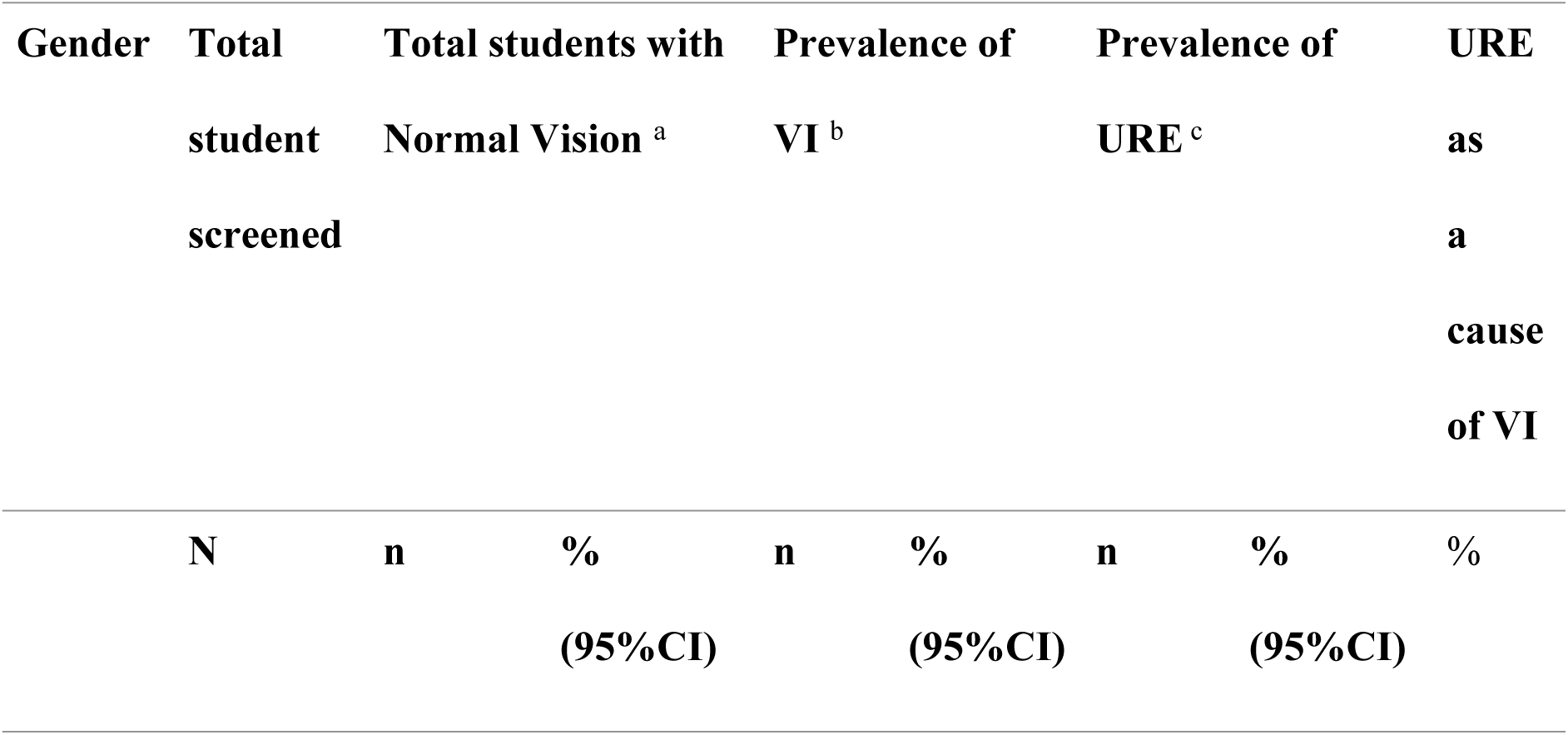
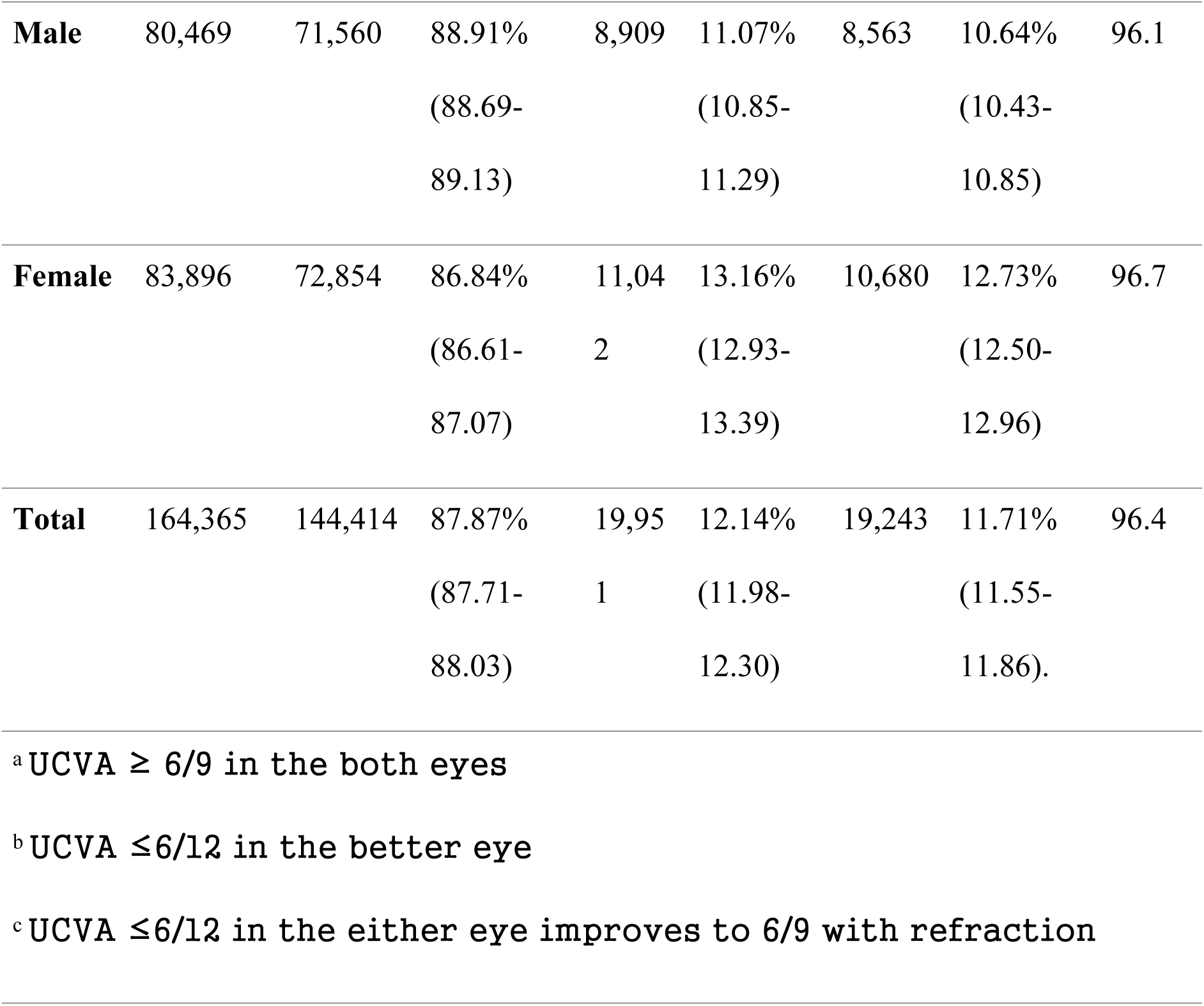
Prevalence of visual impairment and uncorrected refractive error among school children in Bhutan.

### Demographic distribution of refractive errors

The prevalence of URE was significantly higher in female students (12.73%) compared to male students (10.64%) (χ² = 243.6, df = 1, p < 0.001). Similarly, students attending private schools exhibited a significantly greater prevalence of URE (16.45%) than those in public schools (11.32%) (χ² = 269.8, df = 1, p < 0.001). A significant difference was also observed between rural and urban settings, with urban students demonstrating a higher prevalence (16.19%) relative to their rural counterparts (9.56%) (χ² = 1532.8, df = 1, p < 0.001).

Additionally, prevalence varied significantly across regions (χ² = 236.5, df = 2, p < 0.001), with Central (11.78%), East (9.64%), and West (12.68%) regions exhibiting distinct rates, as detailed in **Table 2**.

**Table 2.**
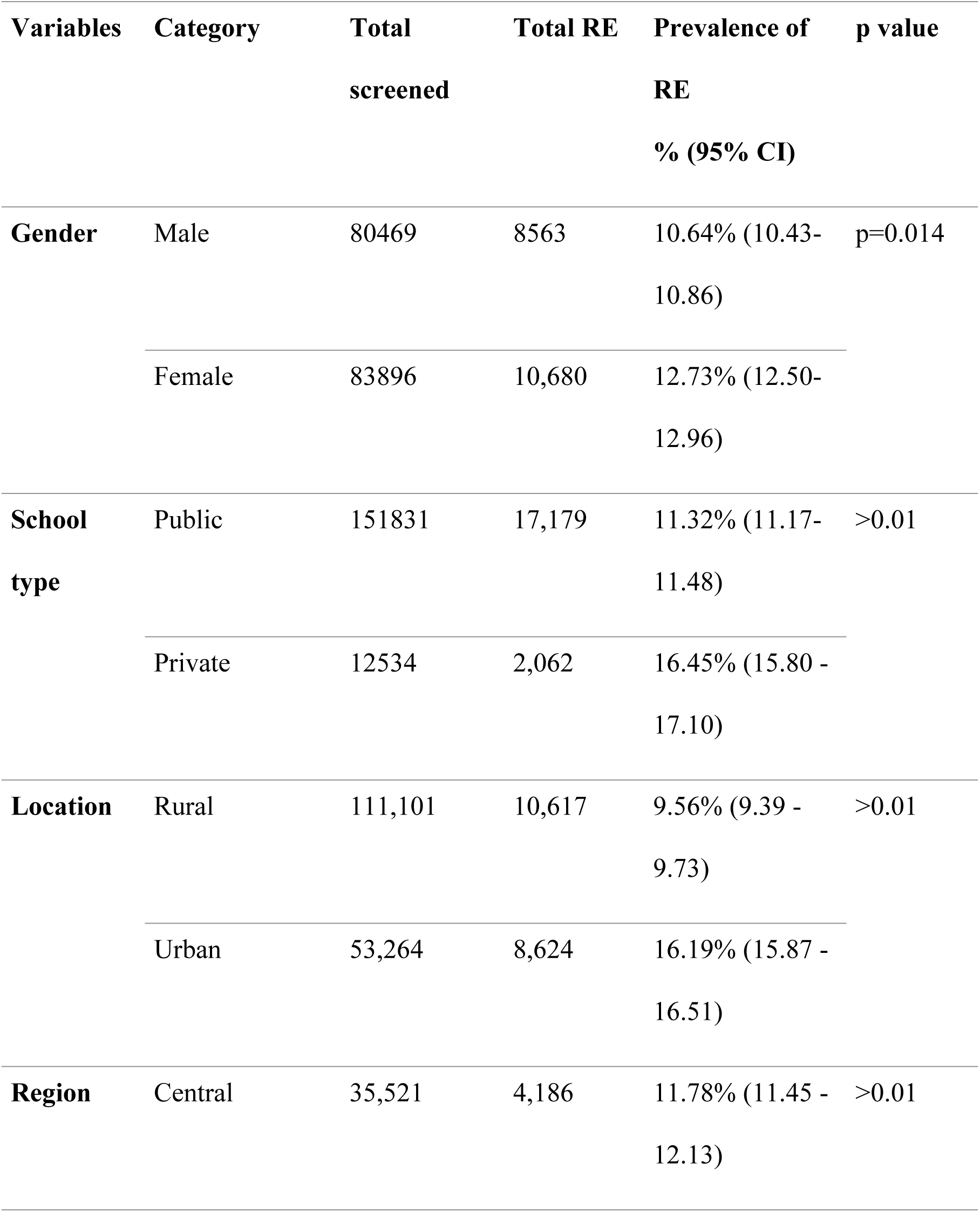
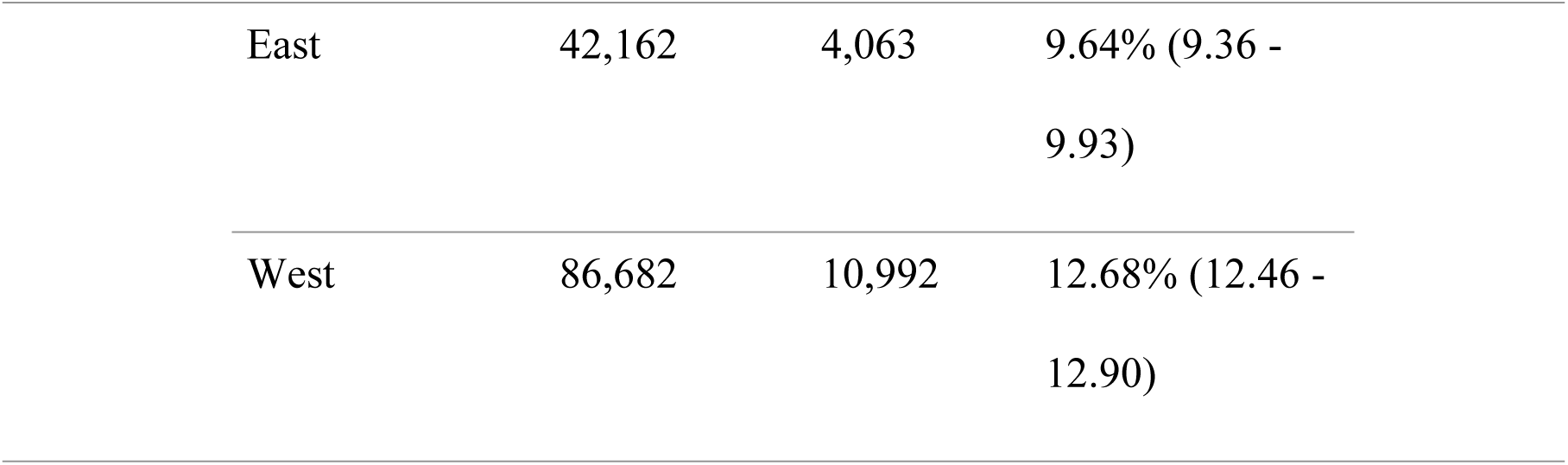
Prevalence of refractive error among school children in Bhutan by gender, school type, location, and region.

| **Variables** | **Category** | **Total**<br>**screened** | **Total RE** | **Prevalence of**<br>**RE**<br>**% (95% CI)** | **p value** |
| --- | --- | --- | --- | --- | --- |
| **Gender** | Male | 80469 | 8563 | 10.64% (10.43-10.86) | p=0.014 |
|  | Female | 83896 | 10,680 | 12.73% (12.50-12.96) |  |
| **School**<br>**type** | Public | 151831 | 17,179 | 11.32% (11.17-11.48) | >0.01 |
|  | Private | 12534 | 2,062 | 16.45% (15.80 -<br>17.10) |  |
| **Location** | Rural | 111,101 | 10,617 | 9.56% (9.39 -<br>9.73) | >0.01 |
|  | Urban | 53,264 | 8,624 | 16.19% (15.87 -<br>16.51) |  |
| **Region** | Central | 35,521 | 4,186 | 11.78% (11.45 -<br>12.13) | >0.01 |

|  |  |  |  |
| --- | --- | --- | --- |
| East | 42,162 | 4,063 | 9.64% (9.36 - 9.93) |
| West | 86,682 | 10,992 | 12.68% (12.46 - 12.90) |

### Geographic Variation

The highest prevalence of RE was observed in Gelephu Thromde at 18.58%, followed by Thimphu Thromde at 17.65% and Phuentsholing Thromde at 15.09%. Conversely, the lowest prevalence rates were reported in Samtse (7.84%), Mongar (8.69%), and Samdrup Jongkhar (8.31%). Screening coverage was complete in all Dzongkhags except for Gasa (76%) and Thimphu Thromde (97.1%). These findings indicate geographic variation in RE burden, with urban Thromdes generally showing higher prevalence rates. District-specific prevalence of URE is presented in **S1Table and Fig 2**, highlighting elevated rates in urban districts such as Gelephu (18.58%), Thimphu Thromde (17.65%), and Phuentsholing Thromde (15.09%), and lower rates in more rural districts such as Samtse (7.84%).

**Fig 2.**
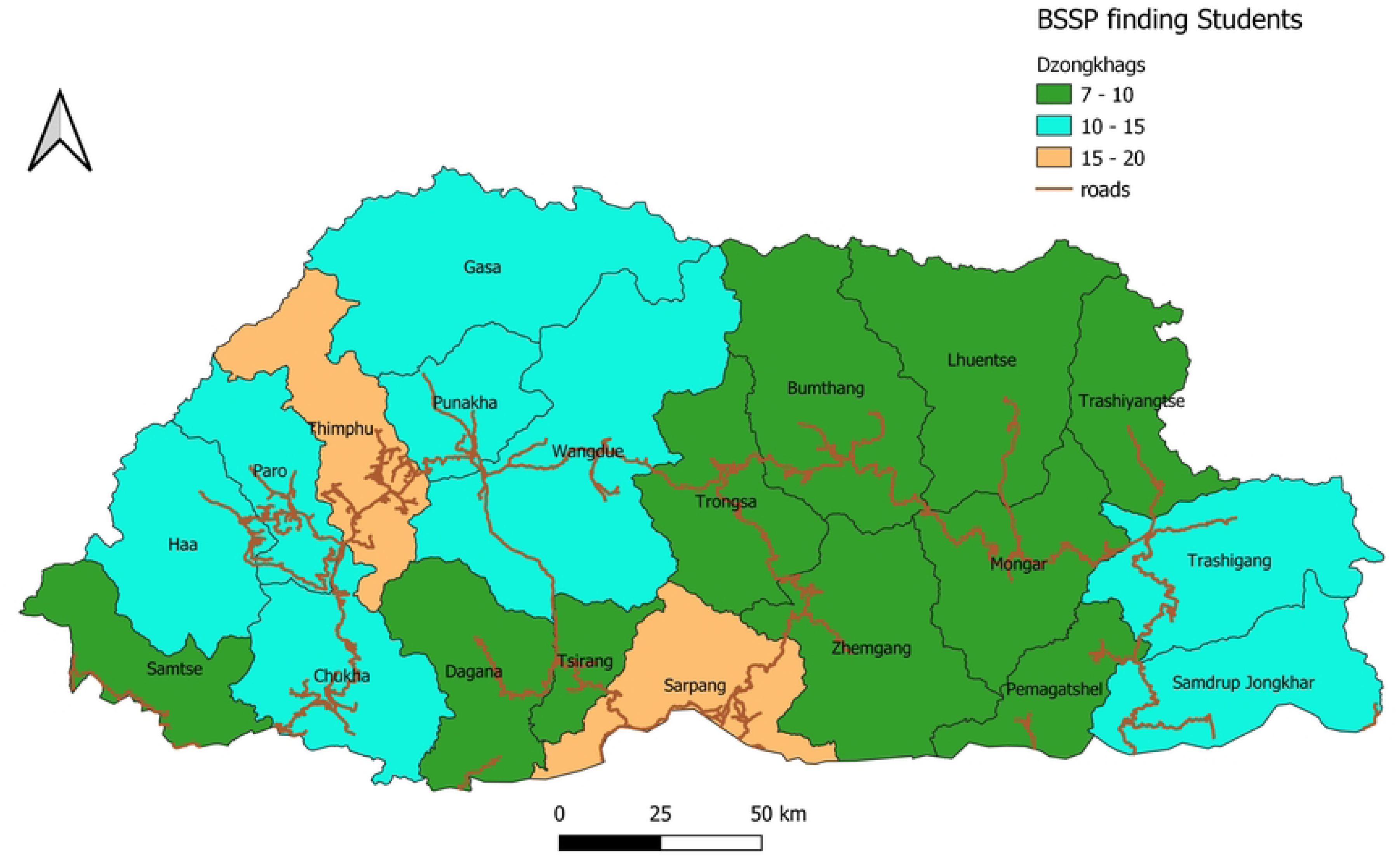
Geographical distribution of prevalence of refractive error among students by districts in Bhutan.

### Predictors of RE

Multivariate regression analysis was conducted to identify significant predictors of RE. Female students (β=3.61, 95% CI: 2.48-4.74, p<0.001) and students enrolled in private schools (β=6.02, 95% CI: 3.40-8.65, p<0.001) exhibited a higher likelihood of RE compared to their respective counterparts. Relative to students attending extended class rooms (ECR), those in lower secondary (β=-5.26, p<0.001), middle secondary (β=-5.10, p=0.001), and primary school (β=-3.20, p=0.001) demonstrated significantly lower RE prevalence. Regional differences did not significantly predict RE. The regression model results for RE are presented in **Table 3**.

**Table 3.** Regression model outputs for refractive error outcome.

| Variable | Category | Coefficient | P-value | 95% CI |  |
| --- | --- | --- | --- | --- | --- |
| Gender | Female vs Male | 3.613 | 0.000 | 2.485 | 4.741 |
| School type | Private vs Public | 6.025 | 0.000 | 3.399 | 8.650 |
| School level | HSS vs ECR | 1.802 | 0.272 | -1.414 | 5.017 |
|  | LSS vs ECR | -5.263 | 0.000 | -7.914 | -2.611 |
|  | MSS vs ECR | -5.096 | 0.001 | -7.966 | -2.226 |
|  | PS vs ECR | -3.199 | 0.001 | -5.027 | -1.370 |
|  | SI vs ECR | 8.798 | 0.078 | -0.990 | 18.586 |
| Region | Eastern vs Central | -1.267 | 0.104 | -2.797 | 0.262 |
|  | Western vs Central | -0.283 | 0.716 | -1.808 | 1.242 |

### Types of Refractive Error

Myopia (n=9,497) accounted for 5.78 % (95% CI: 5.66 to 5.92) of cases, hypermetropia (n=1100) represented 0.67% (95% CI: 0.63-0.71) and astigmatism (n=8644) comprised 5.26% (95% CI: 5.15-5.37). The distribution RE types by demographic characteristics is presented in **Table 4**.

**Table 4.** The distribution of refractive error types by demographics.

| Variable | Category | Myopia | Hypermetropi<br>a | Astigmatism | Total<br>screened |
| --- | --- | --- | --- | --- | --- |
| **Gender** | Male | 3926 | 550 | 3916 | 80469 |
|  | Female | 5571 | 550 | 4728 | 83896 |
| **Location** | Rural | 5328 | 549 | 4740 | 111101 |
|  | Urban | 4169 | 551 | 3904 | 53264 |
| **Public/privat**<br>**e** | Public | 8505 | 997 | 7674 | 151831 |
|  | Private | 989 | 103 | 970 | 12534 |
| **Region** | Central | 2117 | 210 | 1959 | 35,521 |
|  | East | 2051 | 198 | 1814 | 42,162 |
|  | West | 5329 | 692 | 4971 | 86,682 |

Multivariable logistical regression analysis revealed that female students had significantly higher odds of presenting with myopia (OR=1.42) and astigmatism (OR=1.22) compared to male counterparts. Students attending private schools were more likely to exhibit all three types of RE than those in public schools, with the strongest association observed for hypermetropia (OR=1.88). Students residing in rural areas demonstrated slightly lower odds of developing myopia and astigmatism. Relative to the students from the central region, those from the eastern region had lower odds of both myopia (OR=0.86) and astigmatism (OR=0.86); however, regional variations in the prevalence of hypermetropia were not statistically significant.

Females exhibited increased odds of both myopia (OR=1.42) and astigmatism (OR=1.22). Attendance at private schools was associated with significantly higher odds of all three types of RE, with the strongest association observed for hypermetropia (OR=1.88). In contrast, students from rural areas demonstrated slightly lower odds of myopia and astigmatism.

### Spectacle Coverage and Distribution

Of the total number of students diagnosed with RE (n=19234), only 2213 (11.5%) had appropriate spectacles that improved their visual acuity (VA) to 6/9 or better, yielding an effective spectacle coverage of 11.5%. Among the 17,021 students identified as requiring new spectacles, 8595 (50.5%) received ‘ready to clip glasses’ either immediately or within five days following refraction. The remaining 8426 students (49.5%) required customized spectacles to address astigmatism and high-power glasses.

Following follow-up efforts, a total of 7,781 (92.3%) spectacles were successfully delivered to students within four months. Overall, 16,376 children with RE (96.2%) received spectacles. However, 645 children (3.8%) could not be reached, primarily due to school transfers, dropout, or completion of Grade 12.

### Comparison with RESC Study

The BSSS reported a prevalence of VI of 12.14%, whereas the RESC study estimated the prevalence at 16.9%. In both studies, URE emerged as the leading cause of VI, accounting for 94.4% and 96.4% of cases, respectively. Female sex and attendance at urban schools were consistently identified as the primary predictors of RE in both investigations. A comparative summary of the methodologies and key findings of the RESC and BSSS is provided in **Table 6**.

**Table 5:** Predictors of each type of refractive error using multivariable logistic regression.

| Variable | Category | Myopia<br>OR<br>(95%<br>CI) | p-value | Hypermetropia<br>OR (95% CI) | p-value | Astigmatism<br>OR (95%<br>CI) | p-value |
| --- | --- | --- | --- | --- | --- | --- | --- |
| Gender | Female | 1.42<br>(1.36-1.49) | <0.001 | 1.04 (0.91-1.18) | 0.55 | 1.22 (1.17-1.29) | <0.001 |
| Location | Rural | 0.93<br>(0.89-0.97) | 0.001 | 0.95 (0.83-1.09) | 0.48 | 0.96 (0.92-1.00) | 0.05 |
| **School Type** | Private | 1.41<br>(1.31-1.52) | <0.001 | 1.88 (1.51-2.34) | <0.001 | 1.64 (1.53-1.77) | <0.001 |
| **Region** | East | 0.86<br>(0.80-0.93) | <0.001 | 0.88 (0.72-1.07) | 0.2 | 0.86 (0.80-0.92) | <0.001 |
|  | West | 1.04<br>(0.98-1.11) | 0.21 | 1.04 (0.88-1.23) | 0.66 | 1.04 (0.98-1.10) | 0.2 |

**Table 6.** Comparison of methodology and key findings of RESC and BSSS.

| Components | RESC | BSSS |
| --- | --- | --- |
| Year conducted | March to June 2019 | March to December 2019 |
| Target population | School going children (10-15 years) | All school going children (5-18 years) |
| Sampling method | Cluster sampling of schools across Bhutan | Census method |
| Vision screening protocol | RESC Protocol | RESC Protocol |
| Prevalence of Visual Impairment (VA worse than 6/12 in at least one eye) | 16.9% | 12.14% |
| Prevalence of RE | 16% (94.4% of all VI) | 11.71% (96.4% of all VI) |
| Spectacle Coverage | 11.9% | 11.5% |
| Predictors of RE | Female, urban schooling,<br>greater parental education | Female, urban<br>schooling |

## Discussion

The study provides the first national-level data on the prevalence of RE among school-aged children in Bhutan, using data from the Bhutan School Sight Survey (BSSS) 2019. The BSSS achieved near-universal coverage, screening 586 of 589 schools nationwide (99.5%) and assessing 98.7% of enrolled students, representing 94.3% of children aged 5 to 18 years in Bhutan.

The study found that 11.71% of schoolchildren in Bhutan have RE. This prevalence aligns with the estimated pooled prevalence (EPP) reported for South-East Asia, which ranges from 10% to 15%, and is comparable to Nepal’s EPP of 8.4% (95% CI: 4.8-12.9) [11,12]. The similarity between Bhutan and Nepal may reflect shared geographic, demographic, and socioeconomic characteristics.

### Prevalence and distribution of RE

The prevalence of myopia (5.78%), hypermetropia (0.67%), and astigmatism (5.26%) observed in this study is consistent with estimates reported in Southeast Asia, which are 4.9% for myopia, 2.2% for hypermetropia, and 9.8% for astigmatism [11]. Similarly, the EPP of RE in Nepal is 7.1% for myopia, 1.0% hypermetropia, and 2.2% for astigmatism [12]. The relatively low prevalence of hypermetropia, despite the use of cycloplegic refraction, mirrors findings from most Asian studies and may be attributable to the RE definition employed in RESC studies, which excludes mild to moderate hypermetropia (+0.50 to +1.99 D) from prevalence estimates.

The prevalence of myopia in Bhutan is markedly lower than that reported in East Asian countries such as China (36.6%) and South Korea (51.9%) [13,14]. The differences may be attributed to Bhutan’s relatively slower pace of urbanization, higher levels of outdoor activity, relatively lower near-work demands, and limited screen time exposure. However, as Bhutan undergoes socioeconomic transition from a least developed country status, increased rural-to-urban migration, and greater mobile phone penetration, an increase in myopia prevalence is anticipated. Additionally, the recent emergence of ‘left-behind children’ who are primarily cared for by grandparents or relatives while parents seek education or employment abroad, may face increased exposure to unsupervised digital device use, potentially exacerbating the rising trend of myopia [15].

The nearly equivalent prevalence of myopia (5.78%) and astigmatism (5.26%) warrants the need for cycloplegic refraction and comprehensive optical services within school screening programs in Bhutan. However, the prevalence of astigmatism observed in this study is relatively lower than that reported in a study in Xinjiang, China, where astigmatism prevalence exceeded 36% [16]. Studies suggest that ready-made spectacles could address the refractive needs of over 80% of children with URE [17]. However, the implementation of ready-to-clip spectacles during the BSSS encountered challenges like difficulties in maintaining an adequate and diverse stock of lenses and frames in remote screening sites.

### Demographic disparities in RE

The results indicate that gender (female), school type (private), and school location (urban) were significant predictors of RE, consistent with global trends. Female students showed a higher prevalence of URE compared to males. This finding aligns with studies from Asia, which suggests that gender-related disparities in access to eye care and a potentially earlier onset of myopia in females may contribute to the observed differences [18,19].

Students attending private schools exhibited a higher prevalence of RE compared to those in public schools. This difference may be attributed to lifestyle factors, including increased near-work demands and reduced outdoor exposure, both recognized risk factors for RE [20].

Additionally, notable differences in RE prevalence were observed between students in rural (9.56%) and urban (16.19%) schools. The highest prevalence was reported in municipal areas (Thromdes), with the lowest prevalence in districts of eastern Bhutan. These variations may be influenced by urbanization and economic growth. However, a recent meta-analysis of myopia prevalence in Indian school children supports these findings while also indicating a significant increase in prevalence among rural children [21].

### Spectacle coverage and distribution

While the coverage of screening and diagnosis was high, spectacle provision remains a key challenge. At the time of screening, only 11.5% of children with RE were using spectacles, indicating low baseline coverage. Although 96.2% of these children received spectacles within four months, the delay in provision and initial absence of correction reflect gaps in access to refractive services. This highlights the need to strengthen the systems for timely spectacle delivery and follow-up care, components central to the WHO SPECS initiative and the global target to increase eREC by 40% by 2030 [22]. The use of a unique student identification code, provided by the Ministry of Education, facilitated efficient follow-up and spectacle distribution, emphasizing the importance of robust cross-sectoral collaboration to ensure sustainable and timely service delivery.

### Comparison with BSSS

The prevalence of RE found in the BSSS (11.7%) is slightly lower than the 16% reported in the RESC survey conducted in Bhutan [10]. This difference may be due to differences in the age range of the study population; the BSSS included children aged 5–18 years, while the RESC followed the standard protocol targeting children aged 10-15 years. As RE prevalence tends to be higher among older children (12–18 years) compared to younger age groups (5– 11 years), the broader age inclusion in BSSS may have resulted in a lower overall prevalence estimate [19]. Both studies consistently found higher RE prevalence among female students and those in urban areas, indicating persistent demographic patterns in RE distribution. These findings affirm the utility of the RESC methodology for national planning and support the integration of school-based vision services into broader health and education policies.

### Strengths and limitations

A major strength of the study is its near-universal coverage, the use of standardized screening protocols (including cycloplegic refraction), and high participation rates. However, the absence of key variables such as screen time, near work, outdoor activity, parental history, and socioeconomic status limits the ability to comprehensively assess risk factors and design targeted interventions. Additionally, the study did not evaluate compliance with spectacle wear or long-term visual outcomes, which are essential for assessing the overall effectiveness and impact of the program.

### Future research

Future studies should adopt longitudinal study designs to monitor the progression of RE and evaluate the outcomes of interventions over time. Additionally, the inclusion of behavioral and socioeconomic variables is essential to enable a more comprehensive understanding of associated risk factors. Studies assessing the impact, compliance, and acceptability of free spectacle provision are also warranted to inform the effectiveness and sustainability of public health intervention.

### Conclusion

This nationwide analysis confirms that RE is the leading cause of visual impairment (VI) among school-aged children in Bhutan, affecting a little over one in ten students. Although the overall prevalence of RE in Bhutan is relatively lower than in other countries within the region, the disproportionately higher prevalence among female students, those in urban areas, and those attending private schools highlight significant equity gaps. The low baseline spectacle coverage further highlights the disconnect between diagnosis and access to corrective services. Findings from the BSSS demonstrates that large-scale, school-based eye screening programs are both feasible and effective in smaller countries, offering a viable model for national eye health interventions.

As Bhutan advances toward the goal of universal eye health, the findings of this study provide a strategic blueprint for action. Serving as a baseline for RE prevalence among school-aged children, the results have important implications for Bhutan’s national eye health policy and the development of targeted interventions. Addressing RE in children will require substantial investment in follow-up public health measures, timely spectacle provision, and the strengthening of school-based eye health services. With continued efforts and strong cross-sectoral collaboration, Bhutan is well-positioned to lead the region in achieving the WHO 2030 targets for eye health.

## Data Availability

The data used in this study are available upon reasonable request from the Primary Eye Care Program, Jigme Dorji Wangchuck National Referral Hospital, Ministry of Health, Bhutan. Due to ethical restrictions and the data-sharing policy of the Ministry of Health, Bhutan, the dataset cannot be shared publicly. Requests for access can be made by contacting the Ministry of Health, Bhutan at.

## Acknowledgement

The authors thank Dr. Zelalem G Dessie, University of KwaZulu Natal, South Africa for data analysis. The authors also acknowledge the officials of the Policy and Planning Division, and Primary Eye Care Program, Jigme Dorji Wangchuck National Referral Hospital, Ministry of Health, Bhutan for granting the administrative and technical clearance, and sharing data for use in this study.

## Supporting information

**S1Table:** Prevalence of refractive errors among students in all 20 districts (Dzongkhags) and four urban towns (Thromde) in Bhutan

